# Development of the AD*F*ICE_IT clinical decision support system to assist deprescribing of fall-risk increasing drugs: A user-centered design approach

**DOI:** 10.1101/2024.02.06.24301229

**Authors:** Sara S. Groos, Kelly K. de Wildt, Bob van de Loo, Annemiek J. Linn, Stephanie Medlock, Kendrick M. Shaw, Eric K. Herman, Lotta J. Seppala, Kim J. Ploegmakers, Natasja M. van Schoor, Julia C. M. van Weert, Nathalie van der Velde

## Abstract

**Introduction:** Deprescribing fall-risk increasing drugs (FRIDs) is a promising intervention for reducing the risk of falling in older adults. Applying appropriate deprescribing in practice can be difficult due to the outcome uncertainties associated with stopping the use of FRIDs. The AD*F*ICE_IT study addresses this complexity with a clinical decision support system (CDSS) that facilitates optimum deprescribing of FRIDs through use of a fall-risk prediction model, aggregation of deprescribing guidelines, and joint medication management. The development process of the CDSS is described in this paper.

**Methods:** Development followed a user-centered design approach in which users and experts were involved throughout each phase. In phase I, a prototype of the CDSS was developed which involved a scoping and systematic literature review, European survey (*n* = 581), and semi-structured interviews with physicians (*n* = 19), as well as the aggregation and testing of deprescribing guidelines and the development of the fall-risk prediction model. In phase II, the feasibility of the CDSS was tested by means of two usability testing rounds with end users (*n* = 11).

**Results:** The final CDSS consists of five web pages. A connection between the Electronic Health Record allows for the retrieval of patient data into the CDSS. Key design requirements for the CDSS include easy-to-use features for fast-paced clinical practice environments, actionable deprescribing recommendations, information transparency, and visualization of the patient’s fall-risk estimation. Key elements for the software include a modular architecture, open source, and good security.

**Conclusion:** The AD*F*ICE_IT CDSS supports physicians in deprescribing FRIDs optimally to prevent falls in older patients. Due to continuous user and expert involvement, each new feedback round led to an improved version of the system. Currently, a multicenter, cluster-randomized controlled trial with process evaluation at hospitals in the Netherlands is being conducted to test the effect of the CDSS on falls.

## Introduction

Falls and fall-related injuries among older adults are a growing major public health problem [1]. In 2017, 11.7 million older adults in Western Europe requested medical treatment for an injury, of which 8.4 million were fall-related [2]. Injurious falls may result in admission to long term care, loss of independence, reduced mobility, fear of falling, and social isolation, significantly reducing the quality of life for older adults [3–5]. As a result, falls place a significant financial burden on healthcare systems. In Western countries, it is estimated that up to 1.5 percent of the total healthcare expenditures are attributed to fall-related medical care costs [3, 6].

A prominent risk factor for falls in older adults is the use of certain medication classes known as fall-risk increasing drugs (FRIDs). The use or combined use of FRIDs, such as psychotropics and cardiovascular drugs, is associated with adverse effects including orthostatic hypertension, syncope, sedation, and dizziness that can cause accidental falls in older adults [7–10]. Therefore, appropriate deprescribing of FRIDs is recommended for lowering an older adult’s risk of incurring a medication-related fall [11]. Nevertheless, previous studies suggest that deprescribing approaches have yet to be optimized, as current approaches do not sufficiently address the complexity of FRIDs deprescribing for physicians [12, 13].

Deprescribing FRIDs is highly complex due to healthcare professional, patient, cultural and organizational reasons. For example first, physicians themselves may perceive difficulties with *which*, *how* and *when* a FRID or combination of FRIDs should be safely deprescribed. This results in the reluctancy to deprescribe, a phenomena that is especially prominent in the treatment of older patients with polypharmacy and multimorbidity [14, 15]. Second, the unique risk profiles of these patients can greatly increase the complexity of FRIDs deprescribing due to the weighing of competing clinical practice guidelines [15, 16]. As a result, physicians may have a tendency to overestimate the expected benefits of medications (i.e., effective treatment of the chronic condition) and underestimate the potential harms (i.e., an injurious fall) in these patients [17]. Third, physicians may also experience deprescribing reluctance from the patient. Research suggests that low medication-related knowledge in geriatric patients, such as the management of medications, can negatively influence appropriate deprescribing [18].

One way to better support physicians in the deprescribing of FRIDs is by means of a clinical decision support system (CDSS). Such a system has the ability to link individual patient characteristics to a computerized clinical knowledge base which uses information from guidelines to generate patient-specific recommendations back to the physician. These recommendations can subsequently be discussed together with the patient [19, 20]. In the context of FRIDs, a CDSS has the potential to support physicians through a structured deprescribing approach by first signaling *which* medications pose a risk to the individual patient, and subsequently advising *how* and *when* to safely deprescribe each medication (i.e., through guideline integration). In turn, the decision to deprescribe certain medications can be discussed together with the patient to foster joint medication management between physicians and patients. Such shared decision-making has been shown to improve medication-related knowledge and medication adherence in older patients, and could thus enhance the effectiveness of fall preventive care in these patients [21].

Previous studies suggest that medication-related CDSS can improve care outcomes for older patients in a variety of contexts (e.g., inappropriate medication use by patients and polypharmacy, inappropriate prescribing by physicians, falls) [22–24]. However, these studies have not used the potential of a CDSS in deprescribing optimally, such as incorporating data-driven methods, like prediction models, in the system’s clinical knowledge base [24]. Prediction models combine data for multiple risk factors in order to calculate the risk of a future outcome, and help inform subsequent decision making [25]. Thus, in the context of deprescribing optimally, such models could serve as an adjunct to decision-making by allowing the physician and patient to weigh the various treatment options on the basis of the patient’s risk of falling within 12 months. Moreover, such estimates of fall risk could help patients be more aware of their risk of falling and as such motivate them to follow the advice recommended by the physician.

Research suggests that the acceptance of CDSSs among physicians is still hindered by a number of barriers, such as insufficient knowledge with system use, time-consuming, alert fatigue and poor integration into workflow [24, 26–28]. A possible explanation for the considerable number of usability barriers is the lack of involvement by physicians during the development of these systems. A user-centered design approach is an iterative method that involves end users of a system in each stage of the development process. Such an approach has been found to enhance the ease of use and usefulness of clinical study tools – the two important determinants that can influence technology adoption by healthcare professionals [29, 30].

The AD*F*ICE_IT (**A**lerting on adverse **D**rug reactions: **F**alls prevention **I**mprovement through developing a **C**omputerized clinical support system: **E**ffectiveness of **I**ndividualized medica**T**ion withdrawal) project will develop and test a novel deprescribing intervention that aims to prevent medication-related falls in older adults by means of a CDSS for physicians and an online portal for patients. In doing so, it employs a user-centered design approach to develop a data-driven CDSS that aims to provide optimal support for physicians during the deprescribing of FRIDs by generating a personalized fall-risk estimate and guideline-based medication advice tailored to the health conditions of each individual patient. The current paper aims to outline how end-user and expert insights were used to inform the development of our CDSS. We also showcase how end-users (i.e., the physicians) were successfully involved in each stage of the development process to increase the system’s acceptance and adoption in its intended clinical care setting later on.

## Materials and Methods

### Ethics Statement

The Medical Ethics Research Committee of the Amsterdam University Medical Center (Amsterdam UMC, location University of Amsterdam; W19_310 # 19.368) declared that the Medical Research Involving Human Subjects Act did not apply to this study. All study participants gave written informed consent prior to data collection.

### Medical Research Council framework

The overall AD*F*ICE_IT intervention is developed and evaluated following the four phases of the Medical Research Council framework (MRC): (I) the development, (II) the feasibility, (III) the implementation, and (IV) the evaluation phase. The MRC is a guiding theoretical framework for developing, pilot-testing, implementing and evaluating complex health interventions [31]. This paper summarizes the (I) development and (II) feasibility phases for our CDSS. An overview of the studies conducted in these phases is provided in Table 1.

**Table 1.**
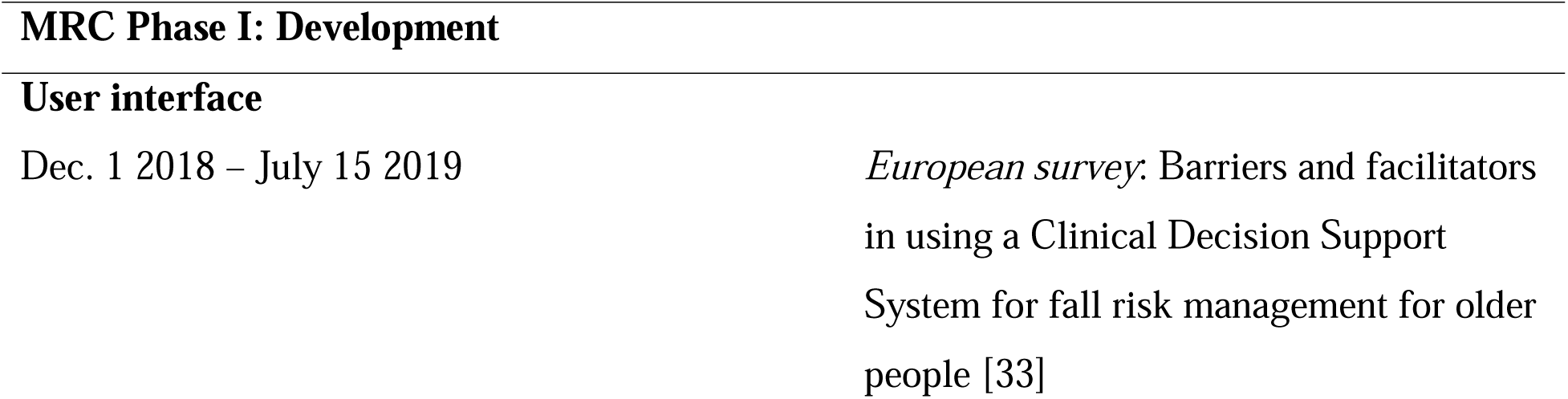
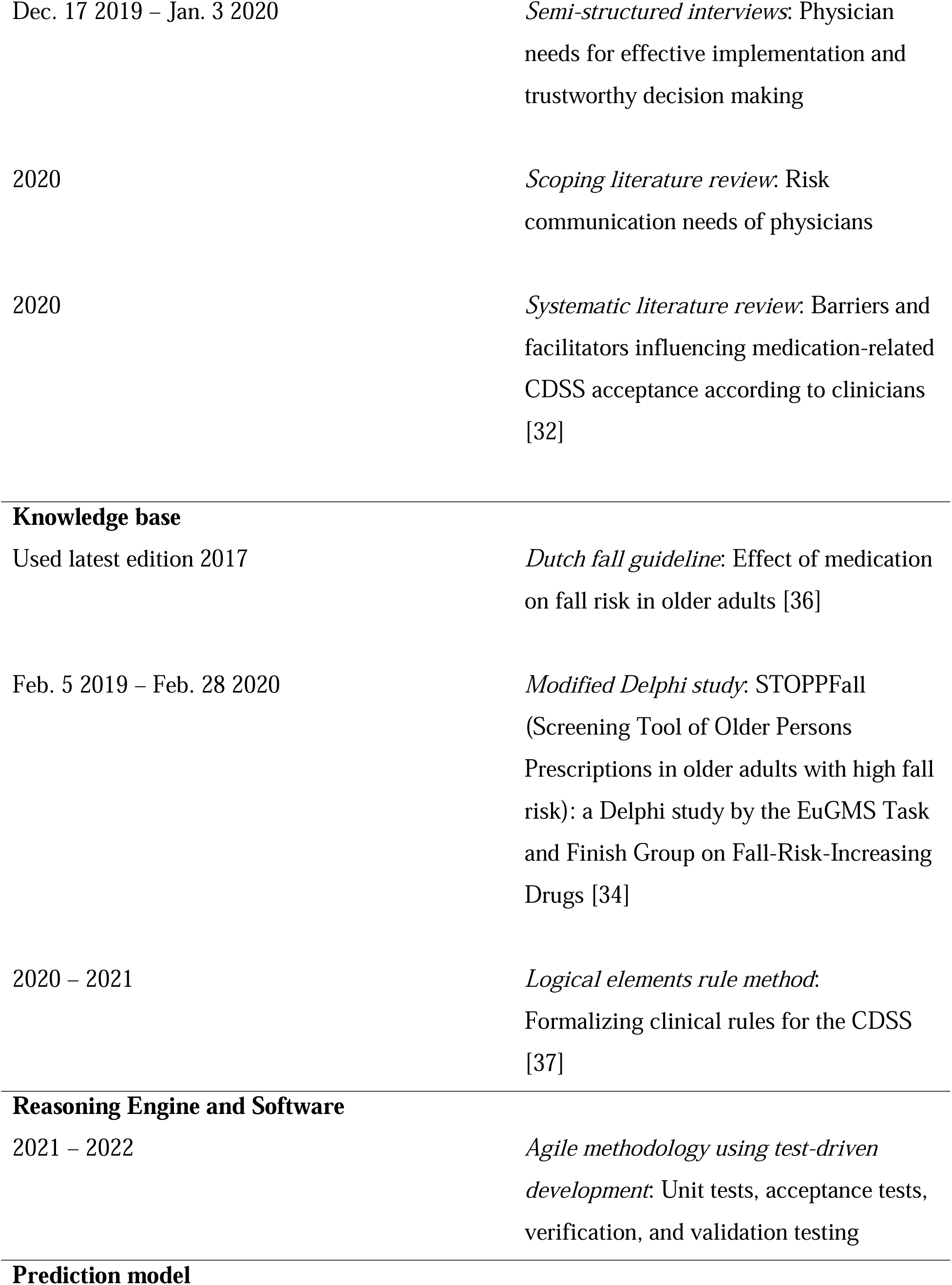
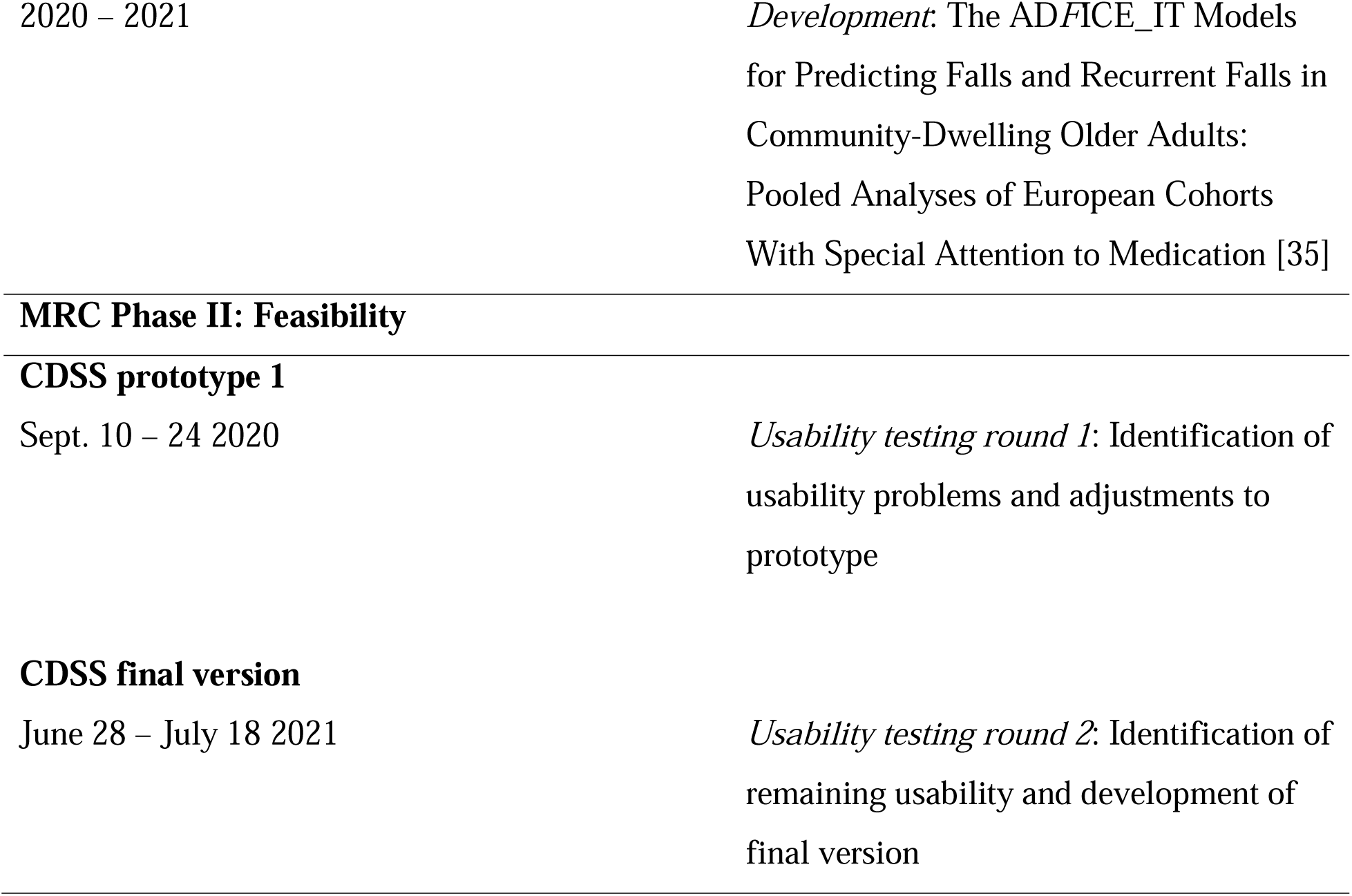
The development process of the CDSS for the AD*F*ICE_IT intervention.

### Phase I: Development

The aim of this phase was to cultivate a robust (theoretical) understanding about *how* to develop the CDSS components of the AD*F*ICE_IT intervention [30], which consisted of the (1) development of the user interface, (2) development of the clinical knowledge base, (3) development of the prediction model, and (4) development of the software. A detailed description of the methods for the systematic literature review, European online survey, modified Delphi study, and development of the fall-risk prediction model have been published elsewhere by the research team [32–35].

#### Development of the user interface

To develop the user interface, a scoping and systematic literature review were carried out, and extended with empirical research by means of a European online survey (*n* = 581), and semi-structured interviews with physicians from Dutch fall clinics (*n* = 19). The scoping literature review assessed the risk communication needs of physicians. The systematic literature review assessed barriers and facilitators to CDSS use [32]. The latter barriers and facilitators to use analysis was expanded on in the survey, which also examined relevant facilitators to use as experienced by physicians [33]. Lastly, semi-structured interviews with physicians assessed the needs for effective implementation and trustworthy decision making. For example, physicians were asked *how* the system should communicate a patient’s fall risk; or *how* the system can best support physicians in making informed decisions about whether or not to deprescribe. The analysis of these studies guided the development of the user interface of the CDSS, such as the system’s functionality, design, and content composition.

#### Development of the knowledge base

The STOPPFall (Screening Tool of Older Persons Prescriptions in older adults with high fall risk) tool [34] and the Dutch fall guideline [36] were used to identify relevant FRIDs in different medication classes. STOPPFall entails a FRIDs list with accompanying guidelines for deprescribing, and was constructed using a modified Delphi technique through consensus effort with 24 panelists from 13 European countries [34]. Both STOPPFall and the Dutch fall guideline form the basis of the system’s clinical knowledge base. To extend the knowledge base specific deprescribing advice for each class of FRIDs was identified from over 30 guidelines, and formalized using an adaptation of the Logical Elements Rule Method (LERM). LERM is a validated method for formalizing clinical rules for decision support (e.g., a CDSS). The method follows a step-by-step approach in which clinical rules are formulated by an informatics knowledge expert, and close collaboration from a clinical expert is sought throughout rule formalization to ensure that the intent of the guidelines are maintained [37]. Specifically, rather than using conjunctive normal form as specified in LERM, the criteria for each rule were formalized as medications to include (e.g., a class of FRIDs), medications to exclude (e.g., non-FRIDs within that class), and conditions (e.g. diagnoses or laboratory values that modify the advice).

#### Development of the reasoning engine and software

Reasoning engine and software development followed an agile methodology, including using test-driven development. Both unit tests (which test specific functions individually) and acceptance tests (which test larger parts of the software as a whole) were used. The software was developed in Node JS, using Express and MariaDB. Jest and TestCafe are used for testing [38–42]. For transparency, all software was developed as open-source software. Experienced developers were involved in creating the software architecture and establishing the development principles. Given the low expected load, clarity of code was prioritized over efficiency and scalability. Importantly, good traceability between the specification for the knowledge base and the implementation of these rules in the software was maintained throughout development, including the provision of evidence behind the recommendations to the end user.

Verification and validation testing were also conducted. Regarding verification, test cases (with specified input and expected output) were developed for each rule in the specification. If any output was not as expected, the corresponding part of the logic was checked for errors and, if needed, corrected. This was repeated until a 100% pass rate was achieved. These tests were added to the test suite and are run every time a change is made to the software. Validation testing took place in two rounds. First, fictional patient cases were constructed based on the specification. Specifically, cases were designed such that each text that the CDSS can produce appeared for at least one patient. The output was reviewed by NvdV to confirm that the advice was clinically sound and reflected the intent of the underlying guidelines. Any identified problems were corrected and the test was repeated. Second, informed consent was obtained from 10 patients and data from these patients was entered into the CDSS. NvdV and an expert in geriatric pharmacy reviewed the cases and commented if they did not agree with the advice. Since clinicians can have differing opinions on the same case, we aimed for 90% agreement with these recommendations.

#### Development of the prediction model

To enable the generation of a patient’s risk of falling within 12 months, a fall-risk prediction model was developed by Van de Loo et al to fit the context of this study using a harmonized dataset (*n* = 5722) of two Dutch cohorts and a German cohort study of community-dwelling older adults (65+) [35]. This prediction model comprises a part of the CDSS’s knowledge base. For background, the outcome variable was defined as any fall (one or more falls) within a one-year follow-up. Candidate predictors were selected based on previously reported risk factors for falls that are easily obtained in clinical practice, such as sociodemographic variables; measures of emotional, cognitive and physical functioning; self-reported chronic conditions; variables related to lifestyle; biomarkers; and use of certain medications. The prediction model was internally validated using an internal-external cross-validation procedure, in which the performance of the model was tested in each of the development cohorts separately following Steyerberg and Harrel [43].

### Phase II: Feasibility

#### Usability testing of the CDSS prototype

The feasibility phase assessed whether the first prototype of the CDSS had any usability problems that needed to be fixed prior to developing the final version of the CDSS. This was achieved through usability testing with geriatric physicians (*n* = 5) from a Dutch academic hospital. Each physician was presented with three hypothetical patient cases that varied in fall risk (e.g., high versus low risk). Each physician completed at least one case. Next, physicians were asked to carry out a scripted navigation consisting of realistic CDSS task scenarios, resembling aspects of clinical documentation (e.g., analyzing a patient’s fall-risk, selecting relevant treatment options, making a referral). Throughout navigation, physicians were prompted to “think aloud” and verbalize their thought process while carrying out the tasks (i.e. using a concurrent think aloud method). Additionally, Camtasia 9, a usability software, was used to record the screen and mouse movements of each physician, including facial expressions and vocalizations (i.e., questions, expressions of confusion) [44].

Usability problems were identified for each physician session, and coded according to the Nielsen usability problems severity rating ranging from *0* = “I don’t agree that this is a usability problem at all” to *4* = “Usability catastrophe: imperative to fix this before product can be released” [45]. The categorization and potential negative impact of each identified usability problem was assessed following the augmented scheme for classifying and prioritizing usability problems [46]. Next, usability problems from all sessions were merged, and for each problem the occurrence of that problem was noted. This led to an overview of usability problems ordered on both severity and occurrence, ranging from the most severe and most often occurring problems to the least severe and least occurring problems. The analysis of the results from this study led to adjustments to the CDSS, resulting in a second version of the prototype. The aforementioned procedure was repeated in a second usability study among six physicians. This allowed us to pinpoint remaining usability issues that needed to be improved prior to the developing the final version of the CDSS.

## Results

The analysis of the results from all studies guided the development of the CDSS. In this article, we report in detail the results of the scoping literature review, semi-structured interviews, software development, and usability testing rounds. The results of the systematic literature review, European online survey, modified Delphi study, and development of the fall-risk prediction model are reported in detail elsewhere by the research team [32–35].

### Phase I: Development

#### Results from the scoping literature review

Regarding the risk communication needs of physicians, the results suggest that physicians prefer a holistic- and patient-specific approach to communicating health-related advice [26]. This approach should include information that is actionable, directive and transparent [47–50]. When visualizing risks, information-orientated graphs were perceived as easier to interpret by physicians, and were found to enhance shared decision-making between physicians and patients [49–51]. Similarly, using a traffic-light coloring system for the presentation of risk is believed to facilitate information processing, which can lead to a more rapid assessment of risk-based information by physicians [47, 49, 51]. Additionally, favorable effects were found on information processing outcomes (e.g., enhanced attention, reduced cognitive load) when the formatting of information and use of terminology was consistent, and when text density and visual clutter was reduced [49, 50].

#### Results from the semi-structured interviews

Results showed that fall risk information should be displayed in color as either a number or percentage. With regard to effective patient-physician communication, a visual graph was viewed as beneficial for the patient, including the ability to print out a patient-friendly handout. This handout should include information about the patient’s personalized fall risk and treatment plan that was discussed during the consultation. Opportunities to read about the benefits and side effects of deprescribing a medication, and having access to additional information (e.g., via hyperlinks) were viewed as important in the decision making to deprescribe. Moreover, information about how a patient’s fall risk is calculated (i.e., the prediction model) was perceived as vital information by physicians that would also enhance the system’s credibility. Lastly, for successful implementation of the CDSS in clinical practice, physicians stressed the importance of reducing completion time (e.g., limiting the amount of mouse clicks).

#### Development of the user interface and knowledge base

Table 2 shows the hierarchical categorization of physicians aggregated system needs from both literature reviews, the European online survey, and the semi-structured interviews with physicians. These key requirements were subsequently operationalized into system features used for the development of the user interface of the first CDSS prototype for usability testing. Regarding the knowledge base of the system, the final set of clinical rules covering 22 classes of FRIDs, with specific deprescribing advice based on diagnoses, lab values, and concurrent medications, including the final fall-risk prediction model were integrated into the CDSS. The final prediction model consisted of the following 14 predictors: educational status, depression, body mass index, grip strength, gait speed, number of functional limitations, systolic blood pressure, at least one fall in the previous 12 months, at least two falls in the previous 12 months, fear of falling, smoking status, use of calcium channel blockers, use of antiepileptics, and use of drugs for urinary frequency and incontinence (see Van de Loo et al. for detailed results) [35].

**Table 2.**
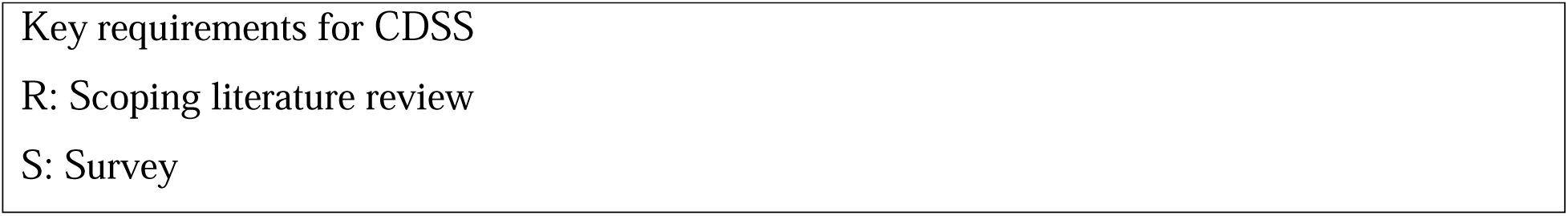
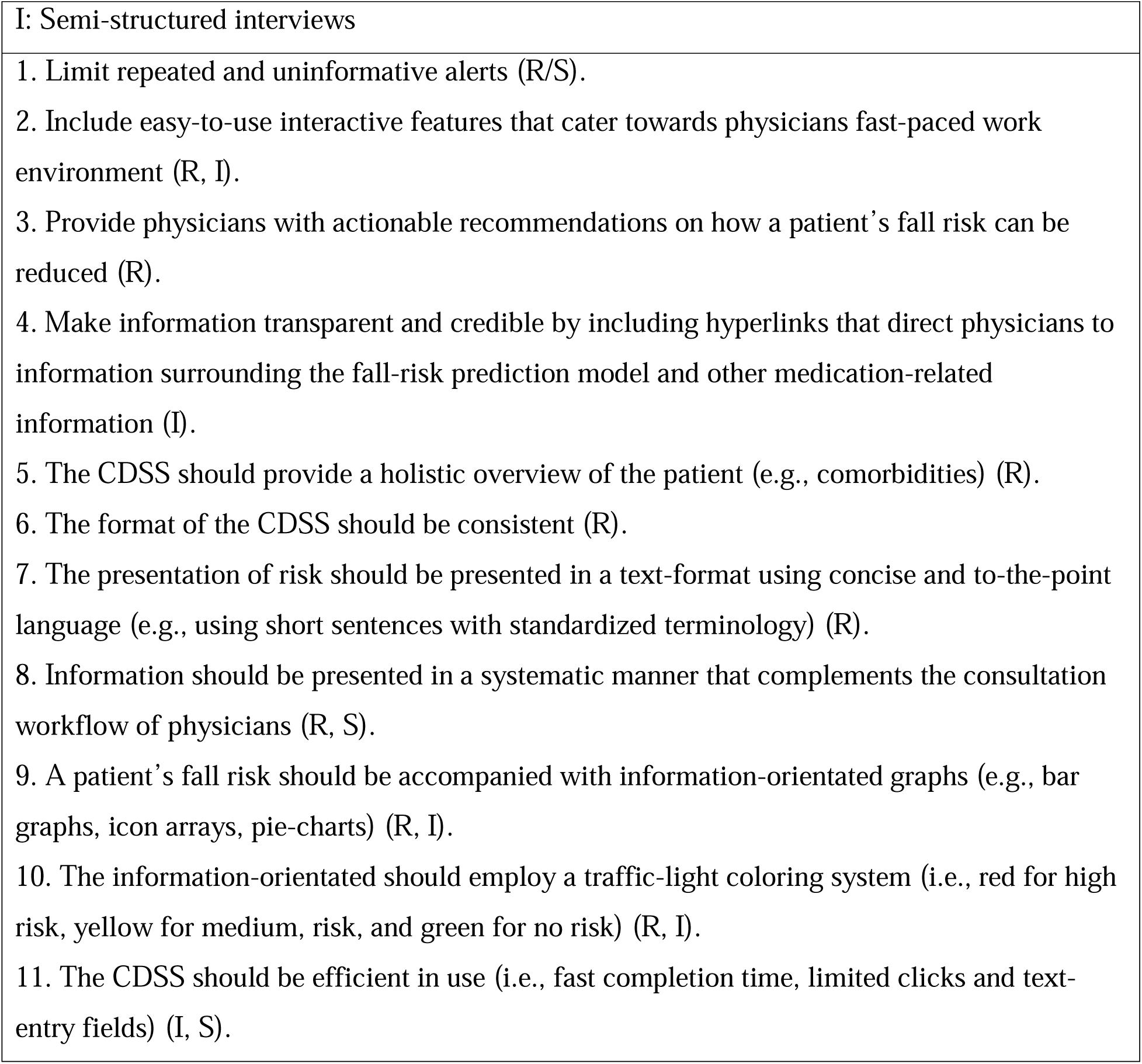
Hierarchical categorization of physicians aggregated needs.

#### Development of the reasoning engine and software (prototype 1)

Following agile development principles, development of the software started by building the smallest part that would be useful, which was the reasoning engine and an interface for verification and validation testing that showed the advice text for the doctor, checkbox options, corresponding patient-friendly text, and references all on one screen. Features were then added in order of priority. This, in combination with test-driven development, lead to a modular architecture with good separation between the connection to the Electronic Health Record (EHR), the logic, and the user interface. This allowed developers to readily modify the interface based on feedback received from phase II. Development as open source software facilitated seeking and incorporating input from external expert developers. Regarding the system’s security, the software is designed to be hosted within the hospital network, and dependencies are minimized to lower the security footprint and improve maintainability. Additionally, the CDSS logic is kept server side to ensure that it cannot be changed from the browser, and that the information seen by the user is always consistent with the saved data. Features were also added to limit data being cached in the user’s browser. Regarding verification and validation testing, after verification testing and the first round of validation testing were completed with no remaining errors detected, the second round of validation testing with real patient data was conducted. This resulted in 100% agreement from one clinician and 95% agreement from the other, which exceeded the minimum threshold of 90% agreement.

### Phase II: Feasibility

#### Results from usability testing rounds

The first usability testing round (*n* = 5 physicians) identified 74 individual usability problems with a mean Nielsen’s severity rating classification of 2.49 (i.e., between minor to major usability problems) [45]. Regarding major to severe usability problems, physicians perceived difficulties with the general navigability of the system (e.g., no “back button”) and the medical terminology used within the system (e.g., what is meant by “lowest dose” or “minimal effective dose” for deprescribing medications). These and similar usability problems were solved and the CDSS underwent a second round of usability testing with 6 physicians. The number of usability problems identified decreased by 16%, and the mean Nielsen’s severity rating was 1.87 (i.e., between cosmetic to minor problems). The most pressing usability problems were improved (e.g., fixing hyperlinks and task-orientated buttons), and the final version of the CDSS was developed. After each usability testing round, the user interface and clinical knowledge base of the CDSS were further optimized, which led to the development of the final CDSS.

#### The final ADFICE_IT CDSS

The source code for the AD*F*ICE_IT software is available at https://github.com/adfice-it. A connection between the EHR and the CDSS was made for the extraction of patient data into the CDSS. This data is used to provide patient-specific advice and to calculate a patient’s personalized fall risk estimate. The final version of the CDSS is depicted in figures 1 through 5. The CDSS consists of 5 web pages. Page 1 (titled “Start” in fig. 1) is the landing page of the CDSS. On this page the physician can (1) check whether relevant patient information (e.g., age, morbidities, list of medications) was correctly extracted from the EHR system, and add missing data for calculating the fall risk estimate; (2) view a graphical representation of the patient’s personalized fall risk estimate by means of a gradient scale, and (3) view a model for shared decision-making that can be implemented during the consultation. Additionally, physicians have access to the patient identifier and personalized fall risk estimate during the entire consultation, as this information is displayed in the form of a horizontal menu bar on all subsequent pages.

**Fig. 1.** Start page of the AD*F*ICE_IT CDSS.

Page 2 (titled “Preparation” in fig. 2) lists each medication taken by the patient and structurally provides the physician with patient-specific deprescribing advice for each listed medication in an attempt to facilitate optimal deprescribing for the physician. Page 2 also provides relevant non-medication related information for preventing falls in older patients (e.g., referral to fall prevention interventions, leaflets, etc.). The clinician can select the treatment options and/or non-medication related information they want to discuss with the patient. Moreover, hyperlinks to third-party sources are provided for additional information about the listed medications (i.e., “Farmacotherapeutisch Kompas”) and related deprescribing advice (i.e., the guidelines used to formulate that advice).

**Fig. 2.** Preparation page of the AD*F*ICE_IT CDSS.

Page 3 (titled “Consult” in fig. 3) provides an overview of the treatment options that the physician selected to discuss together with the patient (i.e., the deprescribing advice or leaflets selected in page 2). Based on the discussion with the patient, a final treatment plan is determined. Page 4 (titled “Advice” in fig. 4) displays the final treatment plan in a patient-friendly format, which is printable by the physician and accessible by the patient via the patient portal. In page 5 (titled “Wrap-up” in fig. 5), the physician is able to copy a summary of the consultation into the patient’s EHR, for future storage.

**Fig. 3.** Consult page of the AD*F*ICE_IT CDSS.

**Fig. 4.** Advice page of the AD*F*ICE_IT CDSS.

**Fig. 5.** Wrap-up page of the AD*F*ICE_IT CDSS.

## Discussion

This study outlined the user-centered development of a CDSS to support physicians in the optimum deprescribing of FRIDs in older adults (65+). The system was developed for the AD*F*ICE_IT project, and intended for use in conjunction with a patient portal. In the development phase (phase I), a robust theoretical understanding about key components of the CDSS was cultivated. For this, both existing and new evidence was relied upon, in which collaboration with end users of the system was sought throughout different stages of development. In phase I, a fall-risk prediction model was developed and internally validated. This prediction model was later integrated into the CDSS. Phase 1 also included the development of the knowledge, reasoning engine and software. Moreover, verification and validation testing were conducted to check for and subsequently correct errors relating to the generation of deprescribing advice. Together, the results from phase I guided the development of the first prototype of the CDSS. In the feasibility phase (phase II), two separate usability testing rounds with geriatric physicians were conducted to identify and address remaining usability issues within the CDSS that could hinder successful implementation of the system later on.

A key strength of this study was the ability to integrate the aggregated deprescribing guidelines and the fall-risk prediction model into the clinical knowledge base of our CDSS. For physicians, deprescribing FRIDs is an often complex task as differences in risk of adverse drug events make it hard to determine whether reducing a particular FRID or combination of FRIDs will result in a significant change in preventing a fall [12, 13]. According to Bloomfield et al., it is exactly this particular complexity in FRIDs deprescribing that has not been effectively addressed in past interventions [12]. This CDSS addresses this need by systematically guiding physicians through the deprescribing of one or more FRIDs. The system does this by first signaling a FRID from a patient’s medication list, and subsequently leverages the stored deprescribing guidelines to advise physicians on how the identified FRID can be deprescribed optimally. Additionally, physicians can leverage the fall-risk prediction model as an adjunct to decision-making by assessing different advice on the basis of the patient’s estimated risk of falling within a 12-month period.

Another key strength of our study was implementing a user-centered design approach to development. This, in our opinion, will greatly influence the acceptance of the system in its intended clinical care setting, as its functionalities and content are tailored to the needs and preferences of its primary end users. Past studies have shown low CDSS acceptance rates and a high number of usability barriers when physicians are not involved in the development process [24, 26–28]. Since the outcomes of the AD*F*ICE_IT intervention are dependent on physicians’ use of the CDSS, insights from the system’s primary end users were gathered, namely physicians treating older patients, at an early stage and throughout the development process of the system, which led to several advantages. For example, the test-driven development allowed us to make changes to the code with confidence that earlier added functionality was not being disrupted.

This also led to a more modular architecture within the code base, which facilitated making the changes suggested by end users during the usability testing rounds. Additionally, while the decision to favor clarity over performance aided in this process, it should be noted that this does carry a limitation whereby the software – in its current form – is not scalable to support hundreds of concurrent users. Moreover, future development could improve testing of the client software further (e.g., unit and acceptance tests) by concentrating manipulation of the display to a few parts of the web page, which could allow for easier testing during development. Lastly, while employing a rigorous verification and validation process led to an improved version of the CDSS after each feedback round, a limitation of this process was the involvement of one of the two clinicians during earlier verification and validation steps. This may have resulted in higher agreement, even though the system did perform well with the second clinician, who was not otherwise involved in the development process.

A final strength of our CDSS is that the system displays several different treatment options (i.e., advice) for each identified FRID (i.e., using guideline-based reasoning), which provides an opportunity for physicians to discuss preferred treatment options together with the patient. This can foster joint medication management, which could help in tackling the observed low rates of deprescribing compliance among patients [52]. In older patients, Van Weert et al. showed that shared decision-making was effective at enhancing both medication-related knowledge and adherence rates, which we believe could have a positive effect on the outcomes of the AD*F*ICE_IT intervention as well [21]. Specifically, the effectiveness of our CDSS (along with the accompanying patient portal) are currently being tested in a multicenter, cluster-randomized controlled trial with process evaluation in several Dutch hospitals.

## Conclusions

This study followed a user-centered design approach to development to develop a CDSS intended for use in fall clinics. The CDSS supports physicians in the optimum deprescribing of FRIDs to prevent falls in older patients (65+), leveraging aggregated deprescribing guidelines, a validated fall-risk prediction model, and shared decision-making. Moreover, due to the continued involvement of end users, we were able to ensure that the CDSS was both useful and user-friendly as each new feedback round led to an improved version of the system. The CDSS is currently being tested in a multicenter, cluster-randomized controlled trial with process evaluation at hospitals in the Netherlands.

## Data Availability

Data from all studies is available upon request. Data from the trial will be made available for other researchers after the study is completed for replication purposes and for original research questions, and is being stored in accordance with General Data Protection Regulation. For this, a request by researchers will need to be submitted for approval from the ADFICE_IT Steering Group. More information about the ADFICE_IT project can be found at http://www.onderzoeknaarvallen.nl. Contact for general queries Contact: Sara S Groos Email address: Address: Amsterdam UMC location University of Amsterdam, Internal Medicine, Section of Geriatric Medicine, Meibergdreef 9, 1105 AZ Amsterdam, Netherlands Contact for scientific queries Contact: Prof Dr. Nathalie van der Velde Address: Amsterdam UMC location University of Amsterdam, Internal Medicine, Section of Geriatric Medicine, Meibergdreef 9, 1105 AZ Amsterdam, Netherlands

## Acknowledgments

The authors thank Tsvetan Yordanov, Rutger Bazen and Stephen Madson for their contributions to the software, and Leonie Westerbeek for her contribution to the user interface. The authors thank Eveline Poelgeest, Gerrit Jan Hafkamp, Irene Gomez Bruinewoud, Hester van der Kroon, Hanna Willems, Suzanne Bleker, Oscar Smeekes and Stephanie van der Woude for their contributions to testing the feasibility of the AD*F*ICE_IT CDSS.

## Availability of materials

Figure translations in English are available upon request from the corresponding author.

